# ANOTHER PERSPECTIVE ON THE RELATIONSHIP BETWEEN BREASTFEEDING, NEWBORN CARE AND MATERNAL PERINATAL DEPRESSION IN A RURAL AFRICAN SETTING

**DOI:** 10.1101/2023.08.26.23294662

**Authors:** Adenike Oluwakemi Ogah, James Aaron Ogbole Oluwasegun Ogah

## Abstract

**Background:** Maternal postpartum depression (PPD) is still one of the major health challenges across the globe with detrimental consequences on newborn bonding, growth and feeding. However, the bidirectional pathways between breastfeeding and PPD have not been sufficiently characterized. This study investigated the risk factors of poor maternal mental health and its relationship with breastfeeding initiation, in a remote community in East Africa.

**Subject and methods:** This was a cross-sectional secondary analysis of the baseline data collected for a 2019 prospective cohort study on infant growth in a rural community in East Africa. Healthy, 529 mother-singleton infant pairs were recruited consecutively from Gitwe district hospital. Maternal peri-partum depression score was obtained using the Edinburgh post-partum depression scale. The cut-off point on the depression scale, used in this study was 6.5.

**Results:** The burden of maternal perinatal depression was 59.7%. Elderly mothers >35years of age (75.9%; p<0.001), poor antenatal visits of less than 4 (74.4%; p<0.001), vaginal delivery (74.7%; p<0.001; OR 5.26; 95% CI 3.57, 7.69), did not give prelacteal feeds (62.4%; p<0.001; OR 7.14; 95% CI 2.94, 16.7), primary education (69.5%, p<0.001), engaged in semi-skilled or skilled jobs (71.8%, p<0.001), poor socioeconomic class (74.8%, p<0.001), Christians (60.9%; p<0.001; OR 8.33; 95% CI 1.89, 33.3), HIV negative (61.6%; p<0.001; OR 4.57; 95% CI 1.90, 11.02), lack of water (82.0%; p<0.001; OR 27.1; 95% CI 16.39, 44.80) and toilet facilities (81.9%; p<0.001; OR 5.68; 95% CI 3.76, 8.58) in their home yard, not being exposed to potentially harmful social habits such as tobacco, alcohol (63.3%; p<0.001; OR 3.22; 95% CI 1.89, 5.56) significantly characterized mothers with high depression score. Peripartum depression was more common among mothers with small for gestational age babies (p=0.004; OR 3.12; 95%CI 1.44, 6.73) and those who initiated breastfeeding early (p<0.001; OR 9.17; 95%CI 5.21, 16.13).

**Conclusion and Recommendations:** The burden of maternal peripartum depression was high in this community and is still on the increase. Therefore, PPD and its risk factors require early recognition, intervention and follow up during the antenatal period up till at least 12 months postpartum, in order to prevent lactation failure and decline in exclusive breastfeeding and continued breastfeeding rates and duration. The poor WASH situation and difficult road access to health facilities in the community requires urgent attention.

## Background

The detrimental impact of perinatal maternal depression (PPD) on the infant, which includes poor breastfeeding, alteration of the infant gut microbial composition and immunity,^1^ growth impairment and underdevelopment have been well documented in literature.^2, 3^ However, the effect of breastfeeding and newborn care on maternal mental health have not been sufficiently investigated. Failure of health workers to understand and address the bidirectional relationship between PPD and breastfeeding have contributed to the increase in lactation failure and decline in the breastfeeding rates and duration in the communities.^4, 5^

Postpartum depression (PPD) is still one of the major maternal health challenges across the globe.^6,7^ PPD is on the rise.^8,9,10^ Estimating the current global PPD prevalence has remained a challenge due to use of varying assessment tools, adoption of different cut-off points for those tools, the varying cultural contexts, and the paucity of related studies in resource-limited environments.^9^

Global mean prevalence of PPD was approximately 15% -25% between 2012 and 2016; 7-30% in 2016;^11^ higher than the 12-13% reported in 2007.^8,9,10^ PPD is estimated to be about 7-20% in high income countries; and could be as high as 45% in low-and middle-income countries.^12^ A systematic review by Atuhaire et al. (2020), yielded a rate of 6.9% -43% PPD in 9 African countries. Uganda had the highest rate of 43% on the EPDS scale, followed by South Africa with 31.7%-39.6%. Morrocco had the least prevalence of PPD at 6.9% to 14%.^13^ Fisher et al, (2012) in their systematic review of 13 papers, covering 17 low and lower-middle income countries, documented an average prevalence of 19.8% (95% CI of 19.5-20.0) for postnatal maternal depression. ^14^ Majority of these studies assessed mothers for PPD, 6 weeks postpartum. This current study, however, aimed to identify PPD much earlier, at birth, to allow prompt intervention. This high burden of maternal perinatal depression in many African countries^14, 15^ have not been matched with adequate mental health systems or human resources for mental health care.^16,17^

Peripartum or postpartum maternal depression may occur within six weeks to six months following childbirth and persist for at least two weeks for such a diagnosis to be made.^13^ Antenatal and peripartum depression can happen even before birth. Depression in mothers before and after birth is characterized by loss of interest in usual events, feelings of sadness, hopelessness and anxiety, sleep challenges, easy fatigability, problems of appetite, and difficulty in coping with daily activities.^18^ As high as 41% of mothers with PPD in a study have entertained thoughts of harming their own infants.^19^

Majority of studies used the Edinburgh postnatal depression scale (EPDS) to diagnose maternal mental health and a cut-off score of ≥9 [with pooled sensitivity of 0.94 (95% CI, 0.68-0.99) and a pooled specificity of 0.77 (95% CI, 0.59-0.88)]. According to Tsai et al.’s systematic review, the sensitivity of EPDS declines, but specificity increases with higher cut-off points above 9.^20^

Fisher et al. identified some risk factors contributing to maternal PPD. These were socioeconomic disadvantage, being younger, being unmarried and giving birth to a female. Protective factors against PPD included having more education and a permanent job.^14^

PPD rate and its impact on newborn feeding and care or vice versa are unknown in Rwanda, the study site of this research. Data pertaining to PPD at birth, its risk factors (maternal, newborn and household factors) and newborn care (birth size and feeding pattern) were obtained to determine the prevalence and risk factors of PPD among these rural mothers; investigate the relationship between PPD, newborn size and breastfeeding initiation.

Since, PPD is a common, preventable and treatable perinatal complication, it is important to increase the PPD awareness of health workers and to encourage them to screen, identify and manage this condition promptly and effectively, to reduce the rate of lactation failure, improve rates of breastfeeding initiation, full exclusive breastfeeding and continued breastfeeding; improved newborn survival and care.

The findings of this study will assist the health care policy makers and providers, in planning tailored interventions to improve maternal mental health all through pregnancy, to better support fetal growth, breastfeeding and the wellbeing of infants globally, especially in the rural areas.

### Concept of the study

Figure 1, shows the factors (maternal, newborn and household) that may contribute to maternal perinatal depression (primary outcome); the relationship between newborn size, feeding (secondary outcomes) and maternal perinatal depression in this study.

**Fig 1:**
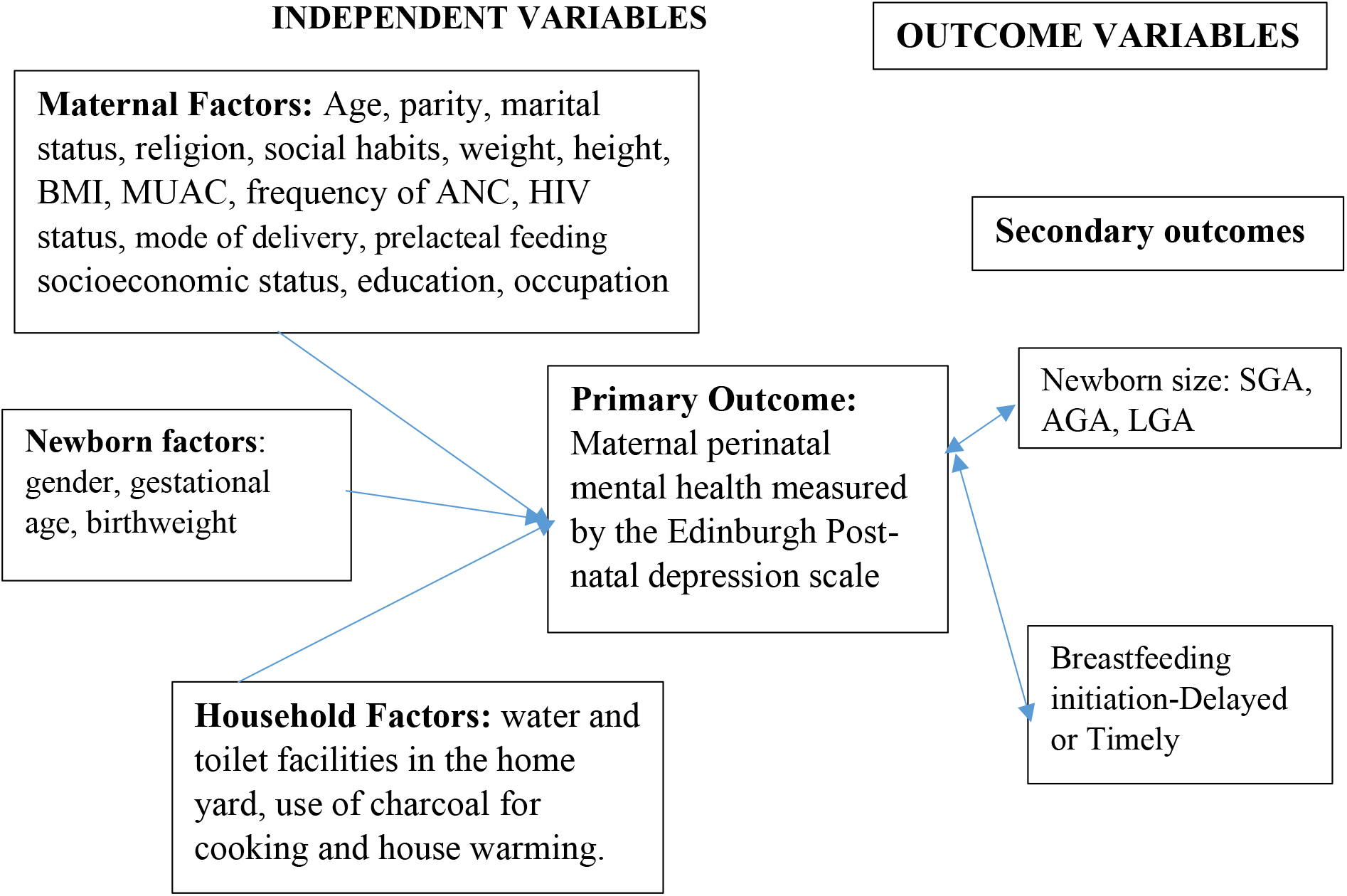
Concept of Perinatal depression and Newborn care.

## Materials and methods

The methods employed in carrying out this study is discussed in this section.

### Study setting

Rwanda is a landlocked country located in central-eastern part of Africa. It is a low-income country with a population of about 13 million people. The country ‘health system is a pyramid with the top of pyramid, being the Ministry of health responsible for sector coordination and oversight, and setting health policies and strategies. Currently, Rwanda has 565 public health facilities; including 504 Health centers; 7 Medicalised Health Centers; 40 District, 4 Provincial, 4 Specialized and 8 Referral hospitals. Health posts are entities working at lowest level and operating under public-private partnership models; there are 1222 Health Posts (HPs) in Rwanda.^13^

Non-availability of regular supplies of clean and safe water, has been a longstanding problem in Rwanda, as a whole, probably because of its landlocked and hilly terrains, making construction and supply of piped water, a major challenge. The public piped water flow infrequently and the taps and pipes may be rusted and breached in some places, especially in the rural areas, further leading to contamination of household water. Many families store rain water in big tanks for use in their homes and this may become polluted (in the writer’s opinion) because of difficulties of cleaning these storage tanks. A few non-profit organizations, such as USAID and Water-for-life have sunk bore holes in strategic locations in a few number of villages accross the country, with the aim of alleviating this water problem.^21^

Gitwe village is located on a high altitude of 1,674 meters above sea level, in the southern province, 240km from Kigali, which is the capital city of Rwanda. Gitwe district hospital began in 1995, immediately after the genocide, for the purposes of providing medical services and later training to this isolated community. The hospital currently has 100% government support, since year 2020. The maximum number of deliveries at the hospital per month was about 200. Some of the challenges in the hospital include poor specialist coverage and few trained health workers, poor supply of equipment, water, electricity, laboratory services and medicines. Challenging cases are referred to the University of Rwanda Teaching Hospital in Butare or Kigali. Gitwe village was selected for this study, because there was no published birth data from this poorly researched, remote community. In 2019, birth depression scores, feeding and growth data on 529 healthy mother-singleton newborn pairs were compiled in this village district hospital delivery and postnatal wards.

### Data source and sample

This was a cross-sectional secondary analysis of the baseline data collected for a 2019 prospective cohort study on infant growth. Mother-newborn pairs were recruited consecutively, on first-come-first-serve basis. Maternal file review and newborn anthropometry [weight (kg), length (cm) and head circumference (cm) measurements, recorded to the nearest decimals] were carried out, soon after birth. Maternal sociodemographic, depression scores, antenatal and nutritional data were obtained using questionnaires, which were read to the mothers and filled by the research assistants. Newborn gestational age and birthweight were estimated. Newborn gestational age was determined using the maternal last menstrual period (LMP), fetal ultrasound gestational age dating (preferably done at first trimester) and/or expanded new Ballard criteria. Newborns were classified into preterm, term and post-term according to their gestational ages; Birthweight regardless of gestational age, with cut-off points of 2.5-3.9kg for normal birth weight.

Newborns were also classified according to their birthweight-gestational age percentiles into small-for-gestational age (SGA), appropriate-for-gestational age (AGA) and large-for-gestational age (LGA): according to WHO using 10^th^-90^th^ percentiles cut-off points for AGA. It is worth noting that comprehensive newborn classification is rarely done in the study site. Time interval from birth to breastfeeding initiation was considered timely, if done within 1 hour and delayed, if done after 1 hour.

To ensure the quality of data collected, 2 registered nurses were trained as research assistants at Gitwe District Hospital on the over-all procedure of mother and newborn anthropometry and data collection by the investigator. The questionnaires were pre-tested before the actual data collection period, on 10 mother-infant pair participants (2% of the total sample). The investigator closely followed the day-to-day data collection process and ensured completeness and consistency of the questionnaires administered each day, before data entry.

### Statistical analysis

Data clean up, cross-checking and coding were done before analysis. These data were entered into Microsoft Excel statistical software for storage and then exported to SPSS version-26 for further analysis. Both descriptive and analytical statistical procedures were utilized. A simple linear regression analysis was used to obtain the threshold cut-off point for maternal depression score. Prevalences of PPD were presented using frequency and percentage tables. Significant risk factors of PPD were identified and associations of PPD with newborn size and time interval to breastfeeding initiation were investigated using Chi test. Odds ratios and 95% confidence intervals were obtained using the logistic regression models (multinomial and binary). Maternal independent variables were age, parity, mode of delivery, marital status, religion, social habits, education, occupation, weight, height, BMI, MUAC, frequency of ANC, HIV status, socioeconomic status. Newborn independent variables were gestational age, birthweight and birthweight-gestational age z-score (WHO). Household independent variables were availability of water and toilet facilities in the home yards. For all, statistical significance was declared at p-value < 0.05. The reporting in this study was guided by the STROBE guidelines for observational studies.^22^

### Ethics

Written permission to collect data was obtained from the Director of Gitwe District Hospital and the eligible mothers gave their informed consent before enrolment. The participants were given research identity numbers and the principal investigator was responsible for the safe keeping of completed questionnaires and collected data, to ensure anonymity and confidentiality of the participants.

## Results

The following are the results obtained from the study.

### Participants

Five hundred and ninety-seven (597) babies were delivered at Gitwe District Hospital in Rwanda, between 3^rd^ January and 9^th^ May 2019, out of which, eligible 529 mother-newborn pairs were enrolled into the study, Figure 2.

**Fig 2:**
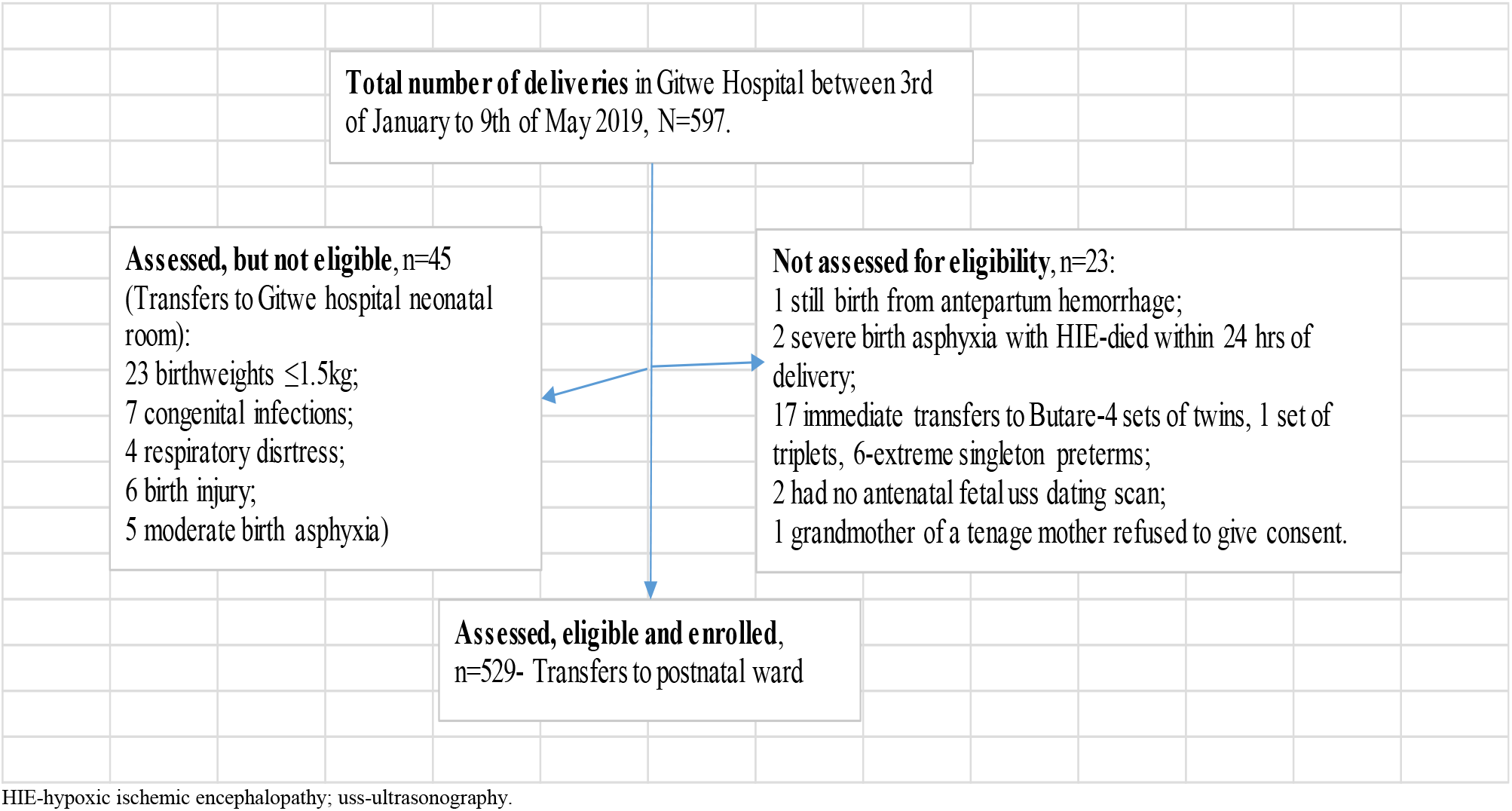
Flow of participants from admission to recruitment into study.

#### Threshold for maternal depression score on the Edinburgh depression scale

A simple linear regression analysis was carried out to determine the maternal depression score at which time interval to initiating breastfeeding from birth was 0 hours. The intercept was 6.391 (rounded up to 6.5) depression score (p<0.001), when time interval to breastfeeding initiation was 0 hours. Hence, all mothers with scores up to 6.5 were coded as 2 (low depression score), while those with scores greater than 6.5 were coded as 1 (high depression score).

The median depression score of these 529 rural women at birth was 7 (IQR 3,9). The percentage of mothers with high depression score was 59.7% (316 out of 529).

#### Significant characteristics of mothers with high depression scores

The significant features of mothers with high depression scores (above 6.5) in this study were: elderly mothers above the age of 35years (60; 75.9%; p<0.001), poor antenatal visits of less than 4 (224; 74.4%; <0.001), vaginal delivery (242; 74.7%; OR 5.26; 95% CI 3.57, 7.69), did not give prelacteal feeds (310; 62.4%; p<0.001; OR 7.14; 95% CI 2.94, 16.7), primary education (242, 69.5%, p<0.001), engaged in semi-skilled or skilled jobs (140, 71.8%, p<0.001), poor socioeconomic class (86, 74.8%, p<0.001), Christians (314, 60.9%, OR 8.33; 95% CI 1.89, 33.3), HIV negative (309, 61.6%; p<0.001; OR 4.57; 95% CI 1.90, 11.02), have no water facility in their home yards (291, 82.0%; p<0.001; OR 27.1; 95% CI 16.39, 44.80), have no toilet facility in their home yards (177, 81.9%; p<0.001; OR 5.68; 95% CI 3.76, 8.58), not exposed to potentially harmful social habits such as tobacco, alcohol (293, 63.3%; p<0.001; OR 3.22; 95% CI 1.89, 5.56) and of fair nutritional status, Table 1. Though, gender of the newborn was not significantly associated with PPD, a higher percentage of mothers with female babies had high depression score, compared to those with male babies (60.2% vs 59.4%). Of note, 3 (0.6%) mothers did not attend antenatal care clinic.

**Table 1:** Features of mothers with High and Low Depression scores, n=529.

| Features | p-value | OR | OR(95%CI) | Depression score |  | Total (100.0) |
| --- | --- | --- | --- | --- | --- | --- |
|  |  |  |  | High (%) | Low (%) |  |
| **Sex of newborn** | 0.852 | 1.03 | 0.73,1.47 |  |  |  |
| Female |  |  |  | 148(60.2) | 98(39.8) | 246 |
| Male |  |  |  | 168(59.4) | 115(40.6) | 283 |
| **Gestational age** | 0.323 |  |  |  |  |  |
| Preterm |  |  |  | 16 (59.3) | 11 (40.7) | 27 |
| Term |  |  |  | 173 (57.1) | 130 (42.9) | 303 |
| Post-term |  |  |  | 127 (63.8) | 72 (36.2) | 199 |
| **Birthweight** | 0.030 |  |  |  |  |  |
| Low birth weight |  |  |  | 34 (60.7) | 22 (39.3) | 56 |
| Normal birth weight |  |  |  | 270 (61.2) | 171 (38.8) | 441 |
| High birth weight |  |  |  | 12 (37.5) | 20 (62.5) | 32 |
| **Prelacteal feeds** | <0.001 | 0.14 | 0.06,0.34 |  |  |  |
| Received |  |  |  | 6 (18.8) | 26 (81.3) | 32 |
| Did not receive |  |  |  | 310 (62.4) | 187 (37.6) | 497 |
| **Age of mother** | 0.001 |  |  |  |  |  |
| Teenage |  |  |  | 14 (60.9) | 9 (39.1) | 23 |
| Very young |  |  |  | 98 (50.3) | 97 (49.7) | 195 |
| Young |  |  |  | 144 (62.1) | 88 (37.9) | 232 |
| Elderly |  |  |  | 60 (75.9) | 19 (24.1) | 79 |
| **Marital status** | 0.597 | 1.18 | 0.64,2.17 |  |  |  |
| Unmarried |  |  |  | 31 (63.3) | 18 (36.7) | 49 |
| Married |  |  |  | 285 (59.4) | 195 (40.6) | 480 |
| **Frequency of antenatal care visits** | <0.001 |  |  |  |  |  |
| Low (<4) |  |  |  | 224 (74.4) | 77 (25.6) | 301 |
| Normal (4-6) |  |  |  | 90 (41.3) | 128 (58.7) | 218 |
| High (>6) |  |  |  | 2 (20.0) | 8 (80.0) | 10 |
| **Mode of delivery** | <0.001 | 0.19 | 0.13,0.28 |  |  |  |
| Cesarean section |  |  |  | 74 (36.1) | 131 (63.9) | 205 |
| Vaginal delivery |  |  |  | 242 (74.7) | 82 (25.3) | 324 |
| **Education** | <0.001 |  |  |  |  |  |
| None/Informal education |  |  |  | 9(34.6) | 17 (65.4) | 26 |
| Primary |  |  |  | 242 (69.5) | 106 (30.5) | 348 |
| 'O' level secondary |  |  |  | 52 (49.5) | 53 (50.5) | 105 |
| 'A' level secondary/College |  |  |  | 10 (31.3) | 22 (68.8) | 32 |
| University |  |  |  | 3(16.7) | 15 (83.3) | 18 |
| **Occupation** | <0.001 |  |  |  |  |  |
| Unemployed/Unskilled |  |  |  | 171 (55.0) | 140 (45.0) | 311 |
| Semi-skilled/Skilled technician |  |  |  | 140 (71.8) | 55 (28.2) | 195 |
| Enterpreneurs |  |  |  | 3 (23.1) | 10 (76.9) | 13 |
| Professional |  |  |  | 2(20.0) | 8 (80.0) | 10 |
| **Exposure to social habits (smokes, alcohol, herbs)** | <0.001 | 0.31 | 0.18,0.53 |  |  |  |
| Yes |  |  |  | 23 (34.8) | 43 (65.2) | 66 |
| No |  |  |  | 293 (63.3) | 170 (36.7) | 463 |
| **Availability of water source in household yard** | <0.001 | 27.1 | 16.39, 44.80 |  |  |  |
| No |  |  |  | 291 (82.0) | 64 (18.0) | 355 |
| Yes |  |  |  | 25 (14.4) | 149 (85.6) | 174 |
| **Availability of toilet facility in household yard** | <0.001 | 5.68 | 3.76, 8.58 |  |  |  |
| No |  |  |  | 177 (81.9) | 39 (18.1) | 216 |
| Yes |  |  |  | 139 (44.4) | 174 (55.6) | 313 |
| **Maternal HIV status** | <0.001 | 0.22 | 0.09,0.53 |  |  |  |
| Positive |  |  |  | 7 (25.9) | 20 (74.1) | 27 |
| Negative |  |  |  | 309 (61.6) | 193 (38.5) | 502 |
| **Maternal Height** | 0.038 | 0.38 | 0.15, 0.98 |  |  |  |
| <150cm |  |  |  | 7 (36.8) | 12 (63.2) | 19 |
| ≥150cm |  |  |  | 309 (60.6) | 201 (39.4) | 510 |
| **BMI (kg/m<sup>2</sup>)** | <0.001 |  |  |  |  |  |
| <18.5 |  |  |  | 1 (25.0) | 3 (75.0) | 4 |
| 18.5-25 |  |  |  | 198 (66.9) | 98 (33.1) | 296 |
| >25 |  |  |  | 117 (51.1) | 112 (48.9) | 229 |
| **Maternal MUAC** | <0.001 | 0.25 | 0.10, 0.60 |  |  |  |
| Low (<23cm) |  |  |  | 7 (28.0) | 18 (72.0) | 25 |
| Not low (≥23cm) |  |  |  | 309 (61.3) | 195 (38.7) | 504 |
| **Religion** | <0.001 | 0.12 | 0.03, 0.53 |  |  |  |
| Moslem |  |  |  | 2 (15.4) | 11(84.6) | 13 |
| Christian |  |  |  | 314 (60.9) | 202 (39.2) | 516 |
| **Parity** | 0.928 |  |  |  |  |  |
| Primiparous (1) |  |  |  | 115 (58.7) | 81 (41.3) | 196 |
| Moderate parity (2-4) |  |  |  | 164 (60.3) | 108 (39.7) | 272 |
| Multiparous (≥5) |  |  |  | 37 (60.7) | 24 (39.3) | 61 |
| **Socioeconomic status** | <0.001 |  |  |  |  |  |
| Destitute/Poor |  |  |  | 86 (74.8) | 29 (25.2) | 115 |
| Middle |  |  |  | 227 (57.2) | 170 (42.8) | 397 |
| Rich |  |  |  | 3 (17.7) | 14 (82.4) | 17 |
| **Total** |  |  |  | **316 (59.7)** | **213 (40.3)** | **529** |

#### Newborn size and maternal depression score

A significantly higher percentage of mothers (74, 23.4%) with high depression score was associated with small for gestational age babies, p=0.011; compared to others with low depression scores (39, 18.3%), Table 2.

**Table 2:** Maternal depression score and newborn size at birth, n=529, p=0.011.

| Maternal depression score | Newborn size |  |  | Total<br>(100.0) |
| --- | --- | --- | --- | --- |
|  | SGA (%) | AGA (%) | LGA (%) |  |
| High score | 74 (23.4) | 228 (72.2) | 14 (4.4) | 316 |
| Low score | 39 (18.3) | 151 (70.9) | 23 (10.8) | 213 |
| **Total** | **113 (21.4)** | **379 (71.6)** | **37 (7.0)** | **529** |

There was significant difference in the median maternal depression scores across the 3 groups of newborns, p=0.015 on the Independent-Samples Kruskal-Wallis Test. Pairwise comparison amongst the 3 groups of newborns, showed that the median maternal depression scores in the SGA group (7, IQR 5,9) and AGA group (7; IQR 3,9) were significantly higher than that of mothers in the large-for-gestational age group (2; IQR 0,8; p=0.011, p=0.046, respectively). However, the median maternal depression scores of the SGA and AGA groups were similar, p= 0.638.

Multinomial logistic regression analysis result showed that the SGA (p=0.004; OR 3.12; 95%CI 1.44, 6.73) and AGA (p=0.010; OR 2.48; 95%CI 1.24, 4.97) babies were more likely to be associated with mothers with high depression scores, compared to the LGA babies.

#### Time interval from birth to breastfeeding initiation and maternal perinatal depression score

The overall median time interval of commencing breastfeeding from birth was 1 hour (IQR 1, 1), range 1-72hours. The median time interval of commencing breastfeeding from birth was 1 hour (IQR 1, 1) for mothers with high depression scores, while it was 1 hour (IQR 1, 2) for mothers with low depression scores. Kruskal-Wallis p-value was <0.001. Likewise, the median maternal depression scores of the mothers in the group that initiated timely breastfeeding was significantly higher (7; IQR 6,9) than those who delayed in initiating breastfeeding (2; IQR 0, 5.25), p<0.001 on Independent-Samples Mann-Whitney U Test.

A higher percentage of mothers with low depression score (73, 34.3%) delayed in initiating breastfeeding, compared with those with high depression score, p<0.001; OR 9.17; 95%CI 5.21, 16.13, Table 3.

**Table 3:** Breastfeeding initiation time interval and maternal depression score, n=529.

| Maternal depression score | Time interval to breastfeeding initiation |  | Total<br>(100.0) |
| --- | --- | --- | --- |
|  | Delayed (%) | Timely |  |
| High score | 17 (5.4) | 299 (94.6) | 316 |
| Low score | 73 (34.3) | 140 (65.7) | 213 |
| **Total** | **90 (17.0)** | **439 (83.0)** | **529** |

## Discussion

The burden of maternal perinatal depression was high at 59.7%, (suggesting progressive increase in the prevalence of PPD), using a threshold point of 6.5 on the EPDS, in this study. Since, the authors did not determine the duration of the symptoms of depression, during data collection, it is difficult to differentiate PPD from ‘baby blues’, which is a milder presentation of emotional disturbance, more frequent and of shorter duration (usually 10 days) than PPD.^23, 24^ This high rate of PPD was probably due to increased exposure to stressful life events in a remote, resource limited setting during pregnancy, delivery and postnatally.^1^ Being involved in demanding semi-skilled or skilled jobs with irregular income, even during pregnancy, and uncertain benefit of paid maternity leave, poor socioeconomic class, and lacking the convenience of water and toilet facilities in their home yards were some of the significant contributors to PPD observed in this study. The lack of nearby water and toilet facilities and the stress of having to leave their homes to fetch water, endangers the hygienic situation in their homes; that could increase the risk of recurrent infections in the community.^25^ This problem of PPD and its risk factors, often remains undiagnosed in routine clinical practice.

Breastfeeding confers many health benefits for mothers and their infants. As such WHO recommends early breastfeeding and breastmilk as the sole food of the infant in the first 6 months of life. Breastfeeding has been known to be associated with less stress or anxiety, promoting relaxation, protecting maternal psychoneuroimmunological status as a result of direct effect of breastfeeding or the release of oxytocin and prolactin during breastfeeding. Oxytocin and Prolactin are known to have mood ameliorating affects.^26,27,28^ Lactation has also been found to reduce levels of stress hormones, especially cortisol. Gonadal, placental and lactogenic hormones are also involved in this interplay between breastfeeding and lower risk of PPD.^29,30^

In contrast to many previous studies,^31^ early breastfeeding initiation was significantly associated with PPD in this current study. Few studies^32,33,34^ have actually suggested that breastfeeding mothers have a higher risk of depression, similar to the findings of this current study. Furthermore, Pippins et al. found that women with prenatal depressive symptoms were not less likely to initiate breastfeeding.^35^ The discrepancies in these study reports may likely be as a result of the interactions between the numerous and complex physiological, psychological, and sociocultural mechanisms and the different study methodology employed.^36^

Early breastfeeding difficulties, negative breastfeeding and breastmilk perception and poor breastfeeding self-efficacy or confidence (feeling of dissatisfaction or inadequacy with breastfeeding)^37^ and maternal peripartum depression, re-inforce each other as they interact in a revolvng cycle, in several studies.^38,39^ If this PPD is allowed to persist, it could pose itself as a greater risk for delayed breastfeeding initiation, lactation failure, early cessation of breastfeeding and introduction of less nutritive, inappropriate and harmful infant feeding practices.^40,41^ Association of SGA baby and PPD in this study, may be as a result of complicated pregnancy, increased burden of advanced newborn care, prolonged stay in the hospital and separation of baby from mother during hospitalization in the neonatal unit. It is difficult to ascertain whether maternal PPD preceded or was responsible for the SGA birth, because data on the onset and duration of depressive symptoms were not collected in this study.

Elderly mothers and mothers with lower educational status with their higher risk of pregnancy complications may be the link associating them with PPD in this study. Of note, mothers, who did not attend ANC or who attended poorly were strongly associated with PPD. Poor ANC attendance may be linked to the difficult hilly untarmacked (often flooded roads during rainfalls) roads leading to the health facilities. Therefore, because of the challenge of accessing the health facility, mothers only seek health care services when they have complications.^42^ Counselling and care received during ANC may have contributed to lower the risk of PPD in this study, as expected. Mothers, who were HIV positive were consistently exposed to one-on-one counselling, unlike mothers, who were HIV negative, during ANC and hence, the lower rate of PPD observed in the HIV positive mothers in this study.^41^ Mothers, who deliver vaginally are assumed to be stronger and therefore receive less attention from health workers, compared to those, who deliver by cesarean section (e.g receiving pain management).^25^ Pain relieving and management of other birth-related health problems may contribute to the lower risk of PPD in mothers, who delivered by cesarean section. For cultural reasons and preference for male child, mothers of female babies tend to be depressed. Mothers, who were exposed to tobacco, alcohol were found to be less depressive in this study as, these harmful agents are often used socially, to elevate mood and suppress pain in the community. Christian mothers, with their nuclear family settings, may lack the ample family support that mothers of non-Christian religion often receive from their polygamous or extended family members during pregnancy and delivery.

Limitations of the study include the lack of history taking on the onset and duration of symptoms of PPD data collection, hence, the author could not determine the temporal relationship between PPD at birth and the secondary outcomes of SGA and delayed breastfeeding initiation. The naturalistic nature of this study made it difficult to determine a causal relationship between breastfeeding and PPD. It may be difficult to compare results of this study with the other similar ones, because of the use of a lower threshold EPDS score.

## Conclusion and Recommendations

Mothers attending antenatal clinics should be screened for risk factors of maternal PPD and PPD, treated and followed up till 12 months postpartum. Early intervention to reduce the prevalence of PPD is highly recommended, in-order to curtail the risk of harm to the infant, rising rate of lactation failure; incomplete exclusive breastfeeding and continued breastfeeding. Repair of access roads to the health facilities, increase campaign for ANC attendance among mothers and the improvement of the WASH situation in the community will significantly reduce the risk of maternal PPD in the community. These interventions are likely to improve infant safety, survival and wellbeing in the community.

## Data Availability

All data produced in the present study are available upon reasonable request to the authors

## Author Contributions

The corresponding author (Dr Ogah Adenike Oluwakemi) conceived and designed the study, collected data, conducted data analysis, interpreted the results, and drafted the manuscript. James Aaron Ogbole Oluwasegun reviewed and edited the manuscript for intellectual content. All the authors approved the final manuscript for submission.

## Acknowledgements

The authors are extremely grateful to the participants involved in this study, to the staff of Gitwe Hospital and clinic in Rwanda and to the research team.

